# Publication performance and trends in dental anxiety research: A comprehensive bibliometric analysis

**DOI:** 10.1101/2025.06.18.25329900

**Authors:** Yuh-Shan Ho, Robert Vautard, Robert Schibbye, Nikolaos Christidis

## Abstract

**Objective:** This bibliometric analysis aimed to systematically evaluate publication performance and identify evolving trends in dental anxiety research over the past three decades, providing a structured overview of key research topics and thematic progression.

**Methods:** The study analyzed 1,556 articles indexed in the Science Citation Index Expanded database from 1991 to 2024. Data extraction included titles, abstracts, author keywords, and *Keywords Plus*. The analytical approach incorporated bibliometric indicators such as total citations, citations per publication, and annual publication trends. A word analysis technique identified five major research topics and their evolution across three distinct periods: 1991–2011, 2012–2019, and 2020–2024.

**Results:** Analysis highlighted significant growth in dental anxiety research publications, particularly in recent years, reflecting increased global interest. The study identified five main thematic areas: *Etiology and Risk Factors, Clinical Presentation and Consequences, Prevalence and Assessment Tools, Treatment and Preventive Interventions*, and *Pediatric Dentistry*. The trends indicated a growing emphasis on multidisciplinary approaches, like cognitive-behavioral therapy and adjunctive therapies such as virtual reality and aromatherapy. Pediatric dentistry consistently emerged as a critical field, underscoring the importance of early interventions.

**Conclusion:** This bibliometric review demonstrated substantial advancements in understanding dental anxiety, emphasizing multidisciplinary treatments and tailored pediatric interventions. Future research should focus on integrating novel therapeutic strategies and refining preventive measures to mitigate dental anxiety effectively.

## Introduction

Dental anxiety is a prevalent yet often underrecognized phenomenon that affects individuals across the lifespan, exerting significant influence on both oral health outcomes and general well-being. ^1,2^ Despite its widespread impact, the term *dental anxiety* is variably defined and encompasses a broad spectrum of emotional and behavioral responses. *Dental phobia*, however, is a formally recognized medical diagnosis with established diagnostic criteria characterized by marked and persistent fear related to dental settings that significantly impairs functioning. Despite modern advances in dental care, fear and anxiety associated with dental treatment persist as significant barriers to regular dental visits, often leading to avoidance behaviors, compromised oral health, and a diminished quality of life. ^3,4^ Recognizing the complexity and variability of dental anxiety is essential for delivering appropriate, individualized treatment, preventing the development of anxiety, and ultimately improving patient health outcomes.

Dental anxiety manifests differently across the lifespan. Among children, dental anxiety is associated with uncertainty and lack of familiarity with dental procedures, environments, or staff. Young children typically fear the unknown, but this fear frequently subsides with age, experience, and increased cognitive understanding of dental routines. ^5,6^ Conversely, persistent dental anxiety in adults is linked to previous traumatic dental experiences, negative interpersonal interactions within dental or healthcare settings, or other unrelated psychological traumas. ^7,8^ Such adults frequently demonstrate pronounced avoidance behaviors, often resulting in severe oral health deterioration. ^9^ Among older adults, cognitive impairments, sensory deficits, and broader health-related anxiety can further complicate dental fear management. In this population, anxiety may be exacerbated by reduced cognitive resilience, sensory hypersensitivity, or misunderstandings about treatment procedures. ^10^

Effective management of dental anxiety requires age-appropriate and individually tailored approaches. In pediatric populations, dental behavior strategies emphasize building trust and familiarity through management techniques such as tell-show-do, positive reinforcement, and parental involvement. ^5,11^ Adolescents and adults with high dental anxiety and dental phobia benefit from psychological interventions, including exposure-based cognitive-behavioral therapy (CBT), systematic desensitization, relaxation techniques, and pharmacological aids such as sedation. ^3,12^ Older patients, particularly those with cognitive decline, may require additional supportive measures, clear communication strategies, and possibly anxiolytic medications under careful supervision to ensure compliance and comfort during treatment. ^10^ An understanding of dental anxiety throughout the lifespan, coupled with the implementation of evidence-based treatment and preventive modalities, is essential to improving oral health outcomes and ensuring patient comfort and compliance.

Bibliometric studies have shown that structured analyses of research trends can guide the development of clinical interventions and prioritize resources effectively, aligning with evolving psychological insights and patient care requirements. ^13,14^ The continuous expansion of scientific literature has significantly advanced our understanding of dental anxiety; however, the sheer volume of available publications creates substantial challenges for clinicians and researchers striving to stay updated with the most relevant and influential findings. ^3,12^ Navigating this extensive literature can feel like searching for a needle in a haystack, as vital research can easily become obscured. ^15^

To manage this complexity, bibliometric analyses have emerged as essential tools. By systematically evaluating publication data, bibliometrics provides comprehensive insights into the progression and thematic evolution of dental anxiety research. ^13^ In the context of dental anxiety, bibliometric analysis offers valuable perspectives for developing evidence-based clinical strategies and interventions. ^1^ This analytical method aids in identifying influential studies and emerging trends, guiding clinical practice, and informing research priorities and funding decisions. ^16,17^ Utilizing bibliometric insights allows dental practitioners, researchers, and healthcare policymakers to effectively manage the growing body of scientific literature, focusing their efforts on evidence-based practices and tailored interventions designed to mitigate dental anxiety and improve patient outcomes. ^18,19^

In this context, the present bibliometric study aims to address the existing challenges by analyzing research trends and publication performance specifically in the field of dental anxiety. The purpose of this bibliometric analysis is not only to conduct a comprehensive citation performance evaluation but also to employ innovative bibliometric methods. Specifically, the analysis incorporates detailed evaluations of article titles, author keywords, *Keywords Plus*, ^20^ and abstracts, ^21^ providing a richer and more nuanced understanding of research trajectories and influential contributions in dental anxiety.

## Methods

The data utilized in this study were obtained from the Web of Science Core Collection (WoSCC) provided by Clarivate Analytics, specifically from the online version of the Science Citation Index Expanded (SCI-EXPANDED). The dataset was last updated on May 1, 2025. Boolean operators such as OR and AND were employed when formulating the search queries. To ensure accurate retrieval, multi-word terms were enclosed in quotation marks (“”). The OR operator was used to identify records that contained at least one of the specified keywords within the Topic field, which encompasses the title, abstract, author keywords, and *Keywords Plus*.

The search was conducted using targeted keywords, including “dental anxiety”, “dental fear”, and “dental phobia”. To have more accurate analysis results, uncommon terms ^22^: “fear of dental”, “anxiety in dental”, “anxiety during dental”, “dental fears”, “fear dental”, “dental fear anxiety”, “dental trait anxiety”, “fearful dental”, “dentally fearful”, “odontophobia”, “anxiety among dental”, “anxiety about dental”, “dentophobia”, “fear of dentistry”, “fear of dentists”, “anxiety in dentistry”, “anxiety on dental”, “odontophobics”, “anxiety before dental”, “fear during dental”, “anxiety of dental”, “anxiety toward dental”, “anxiety towards dental”, “dental anxieties”, “dental care anxiety”, “dental care fear”, “dental patient anxiety”, “dental treatment fear”, “dentist fear”, “dentophobic”, “phobia and dental”, “anxiety regarding dental”, “dental anxiety phobia”, “dental patients’ anxiety”, “dental phobias”, “dentist phobia”, “dentists’ anxiety”, “dentists’ fear”, “fear about dental”, “fear for dental”, “fear of dentist”, “fearful of dental”, “fears about dental”, “fears in dental”, “odontophobic”, “phobia of dental”, “anxiety from dental”, “anxiety with dental”, “dental state anxiety”, “dental treatment phobia”, “dentophobics”, “feared by dental”, “anxieties about dental”, “anxiety about dentistry”, “anxiety among dentistry”, “anxiety around dental”, “anxiety following dental”, “anxiety on dentists”, “anxiety towards dentists”, “anxiety-eliciting dental”, “dental and anxiety”, “dental fearful”, “dental hygiene fears”, “dental injection phobia”, “dental procedure anxiety”, “dental state fear”, “dental trait fear”, “dental treatment anxieties”, “dental needle phobia”, “dental-related anxieties”, “dental-related fear”, “dental-specific anxiety”, “dentist anxiety”, “dentist for fear”, “dentists allay fear”, “dentists for fear”, “dentists through phobia”, “fear among dentists”, “fear in dentistry”, “fearful about dentistry”, “fearful of dentists”, “fearing dental”, “fears of dental”, “phobia in dentistry”, “phobias about dentistry”, “phobias within dentistry”, “fear of intra-oral injections”, “intra-oral injection fear”, “intra-oral injection phobia”, “specific phobia dentist”, and “specific phobia dentistry” in SCI-EXPANDED were also considered. A total of 2,073 documents, including 2,066 documents (99.7% of 2,073 documents), containing search keywords in the terms of TOPIC were searched-out in the SCI-EXPANDED database published between 1991 and 2024.

The complete records of documents retrieved from SCI-EXPANDED, including annual citation counts, were downloaded into Microsoft 365 Excel for analysis. Following the methodology previously described by our research group, ^23,24^ manual coding was performed to refine and enrich the dataset. Additionally, journal impact factors (IF2023) were obtained from the 2024 edition of the Journal Citation Reports (JCR).

In 2011, the use of the “front page” – comprising the title, abstract, and author keywords – was introduced as a filtering strategy to improve search precision when using the Topic (*TS*) field in the Web of Science Core Collection for bibliometric research. ^25,26^ This approach aims to concentrate on the most pertinent content by restricting results to key textual components, thereby minimizing the inclusion of irrelevant publications. When applied to medical research topics within SCI-EXPANDED, the front-page filter revealed significant discrepancies, highlighting its effectiveness in enhancing data quality. For example, its implementation resulted in a 4.0% reduction in irrelevant records in dental education, ^27^ a 15% deviation in studies on temporomandibular disorders, ^15^ and a notable 31% deviation in bruxism research. ^28^ The search for documents containing the search keywords in their “front page” yielded a total of 1,884 documents, which accounted for 91% of the 2,066 initially identified documents.

In the WoSCC database, the reprint author is identified as the corresponding author; however, in this study, the term “corresponding author” was used instead. ^29^ For articles with a single author, single institution, and single country without explicitly defined authorship roles, the sole author was designated as both the first and corresponding author, and the institution and country were classified accordingly. ^23^ In instances where multiple corresponding authors were indicated, all listed corresponding authors, along with their respective institutions and countries, were included in the analysis. ^23^ Furthermore, for articles in SCI-EXPANDED that provided only addresses without naming specific affiliations, these addresses were verified and updated to reflect the appropriate institutional affiliations. ^23^

Following the methodology described by Chiu and Ho in 2005, ^30^ affiliations from England, Scotland, Northern Ireland, and Wales were consolidated under the designation of the United Kingdom (UK). Similarly, affiliations listed as Turkiye and New Caledonia were reclassified as Turkey ^31^ and France, ^32^ respectively. Affiliations from Hong Kong were categorized under China. ^26^ Additionally, affiliations originally recorded as Yugoslavia were reviewed and reclassified as either Slovenia or Serbia based on institutional details. ^33^

The evaluation of publications in this study was conducted using three citation indicators:

- *C*_year_: This indicator represents the number of citations received from the WoSCC in a specific year (e.g., *C*_2024_ denotes the citation count for the year 2024) as proposed by Ho in 2012. ^18^
- *TC*_year_: This indicator reflects the total number of citations received from the WoSCC from the year of publication until the end of the most recent year (2024 in this study; denoted as *TC*_2024_), as introduced by Wang et al. in 2011. ^34^
- *CPP*_year_: The average number of citations per publication, calculated as *CPP*_2024_ = *TC*_2024_/*TP*, where *TP* denotes the total number of publications. This measure was suggested by Ho in 2013. ^35^

The citation indicators can be applied for wide range of categories, for example, total and annual publications, as well as publications in a document type, language, Web of Science category, journal, country, institution, author, and an article.

In 2014, six publication indicators were proposed to evaluate the publication performance of countries and institutions ^36,37^ as:

1) *TP*: Total number of articles published.
2) *IP*: Number of articles published by a single country (*IP*_C_) or institution (*IP*_I_).
3) *CP*: Number of internationally collaborative articles (*CP*_C_) or inter-institutionally collaborative articles (*CP*_I_).
4) *FP*: Number of first-author articles.
5) *RP*: Number of corresponding-author articles.
6) *SP*: Number of single-author articles.

Moreover, six citation indicators (*CPP*_2024_) corresponding to these publication indicators were used to evaluate the impact of publications on document types, journals, countries and institutions, as proposed by Ho and Mukul in 2021. ^38^

### Statements

#### Statement about originality

The research conducted in this manuscript is original, not presently under consideration for publication elsewhere, free of conflict of interest and conducted by the highest principles of human subject welfare. The authors alone are responsible for the content and writing of the paper.

#### Ethics statement

Not applicable since this is a bibliometric analysis in already published papers that are publicly available on Web of Science Core Collection

#### Patient consent

Not applicable since this study does not include any participants and does not report any patient data

## Results and Discussion

### Characteristics of document types and languages

To evaluate the characteristics of document types within a specific research field, key bibliometric indicators such as the average number of citations per publication per year (*CPP_year_*) and the average number of authors per publication (*APP*) are commonly employed. ^39^ Between 1991 and 2024, a total of 1,884 documents related to dental anxiety were indexed in the SCI-EXPANDED database, encompassing 11 different document types, as summarized in Table 1. Among these, research articles were the most prevalent, accounting for 1,556 documents (83% of the total), with an average of 4.4 authors per publication. Of all document types, reviews (122 publications) exhibited the highest citation impact, with a *CPP_2024_* of 30, based on 3,683 total citations. This made reviews, on average, 1.4 times more cited than research articles. This citation ratio between reviews and articles is slightly lower than that observed in other dental research domains. For example, in dental education, reviews were cited 1.8 times more frequently than articles, ^27^ while in the fields of temporomandibular disorders and bruxism, the ratio was 1.7. ^15,28^ However, in third molar research, reviews were cited less frequently than articles, with a *CPP_year_* that was only 0.73 times that of articles. ^40^

**Table 1.** Citations and authors according to the document type.

| Document type | *TP* | % | *TP\** | *AU* | *APP* | *TC*<sub>2024</sub> | *CPP*<sub>2024</sub> |
| --- | --- | --- | --- | --- | --- | --- | --- |
| Article | 1,556 | 83 | 1,556 | 6,892 | 4.4 | 31,688 | 20 |
| Meeting abstract | 147 | 7.8 | 147 | 481 | 3.3 | 11 | 0.075 |
| Review | 122 | 6.5 | 122 | 616 | 5.0 | 3,683 | 30 |
| Editorial material | 34 | 1.8 | 32 | 65 | 2.0 | 103 | 3.0 |
| Proceedings paper | 17 | 0.90 | 17 | 74 | 4.4 | 403 | 24 |
| Early access | 14 | 0.74 | 14 | 65 | 4.6 | 27 | 1.9 |
| Letter | 10 | 0.53 | 9 | 14 | 1.6 | 1 | 0.10 |
| News item | 9 | 0.48 | 6 | 6 | 1.0 | 1 | 0.11 |
| Correction | 5 | 0.27 | 5 | 20 | 4.0 | 7 | 1.4 |
| Discussion | 1 | 0.053 | 1 | 1 | 1.0 | 0 | 0 |
| Retracted publication | 1 | 0.053 | 1 | 10 | 10 | 22 | 22 |

*TP*: total number of publications; *TP\**: total number of publications with author information in the SCI-EXPANDED database; %: percentage of articles in all articles; *AU*: total number of authors; *APP*: average number of authors per publication; *TC*<sub>2024</sub>: total number of citations from WoSCC since publication year until the end of 2024; *CPP*<sub>2024</sub>: average number of citations per publication (*TC*<sub>2024</sub>/*TP*); N/A: not available.

Within the corpus of 1,884 dental anxiety-related publications, 45 were identified as highly cited papers, defined as having a total citation count (*TC_2024_*) of 100 or more. ^41^ This group comprised 37 articles and 8 reviews.

It should be noted that documents in the WoSCC can be assigned to multiple document types. For example, 39 proceedings papers, 10 retracted publications, and one early access paper were all simultaneously classified as articles. Consequently, the cumulative percentages in Table 1 exceed 100%. ^42^

Different document types contribute to bibliometric analyses in distinct ways. Among them, research articles – typically structured with sections including introduction, methods, results, discussion, and conclusion – are generally regarded as the most significant for evaluating scholarly output in a given field. ^43^ In the case of dental anxiety research, a total of 1,556 articles were published across four languages. English was overwhelmingly dominant, representing 1,550 articles (99.6%), followed by German (4 articles), and one article each in Croatian and Dutch.

### Characteristics of publication outputs

To gain a deeper understanding of the citation lifespan of research articles, Chuang et al. in 2007 ^44^ proposed analyzing the relationship between the average number of citations per article in a given year (as indexed in WoSCC) and the age of the article, defined as the number of years since publication. Figure 1 presents a comparative analysis across various dental-related research topics. In the field of dental anxiety, 1,556 articles received a total of 550 citations in their publication year (*C_0_* = 550), yielding an average of 0.353 citations per publication (*CPP_0_* = 0.353). Citation impact peaked in the tenth year after publication, during which 703 articles accumulated 1,595 citations, resulting in the highest *CPP_10_* of 2.27 (1,595/703). This pattern suggests enduring interest in the field, although the peak was slightly lower than that observed in other dental research areas. For example, bruxism exhibited a *CPP_4_* of 2.51 in the fourth full year post-publication, ^28^ temporomandibular disorders reached a *CPP_5_*of 2.49 ^15^, and third molar research had a *CPP_5_* of 2.39. ^40^ In contrast, dental education demonstrated a lower long-term citation impact, with a *CPP*_2_ of only 1.74, ^27^ despite having the highest initial citation impact. Of the 3,483 dental education articles, 2,164 citations were received in the year of publication, resulting in the highest *CPP_0_* across all topics at 0.621. This was followed by bruxism (0.389), dental anxiety (0.353), temporomandibular disorders (0.348), and third molar research (0.300). These findings indicate that while dental education articles attract early attention, their long-term citation impact is comparatively modest. Conversely, articles on dental anxiety demonstrate a more extended citation lifespan, reflecting sustained academic interest.

**Figure 1.**
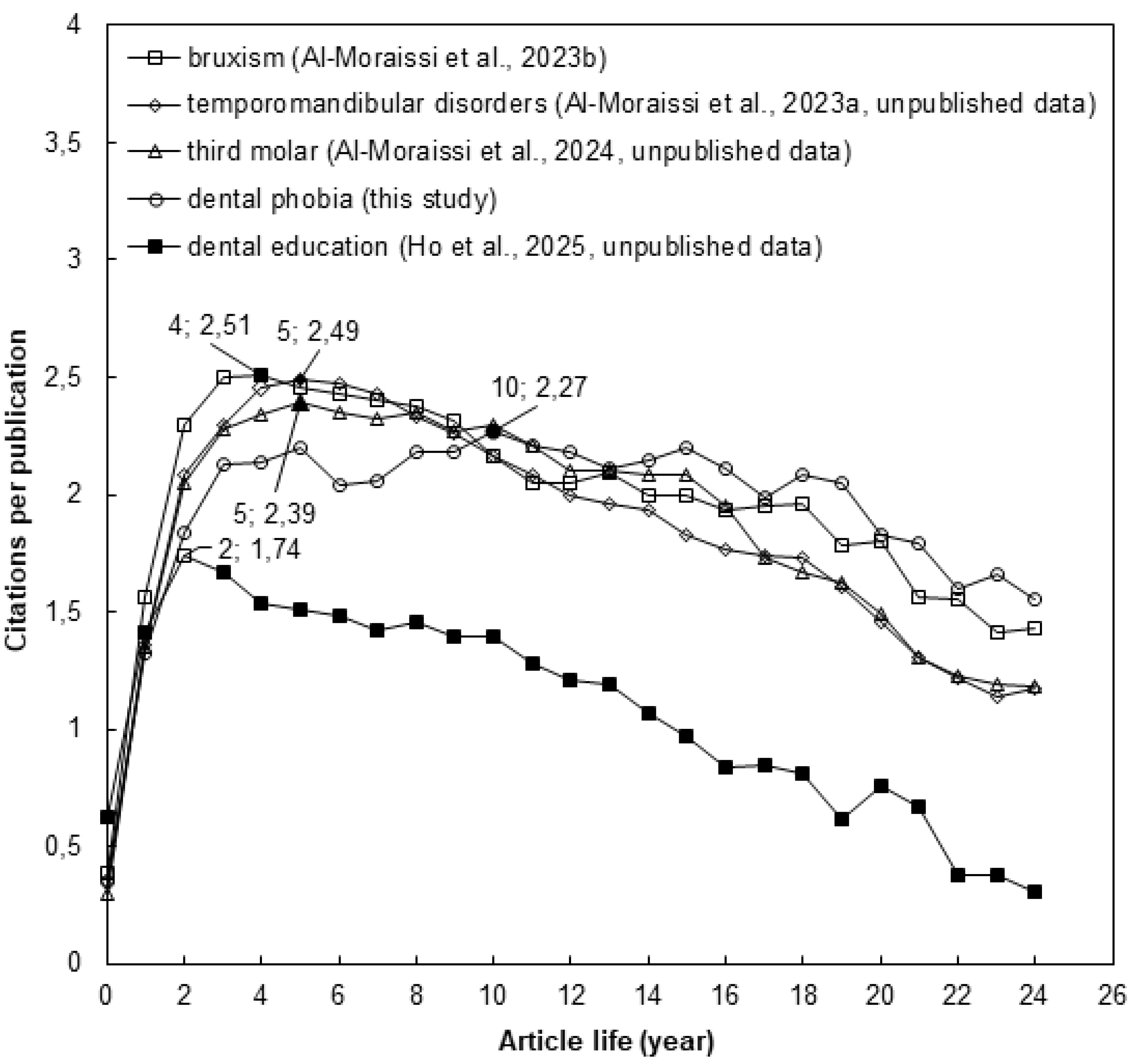
Citations per publication by article life.

To assess cumulative citation impact from the year of publication through the end of 2024, the citation indicator *CPP_2024_* was used. This metric allowed for an exploration of the relationship between *CPP_2024_*and the annual number of publications (*TP*) by year, offering insights into both research impact and publication trends within the topic. ^35^ The annual number of dental anxiety-related articles indexed in SCI-EXPANDED was counted and is illustrated in Figure 1. It is widely recognized that citation accumulation requires time. For example, 128 articles published in 2024 had received a total of 50 citations by the end of the same year, resulting in a *CPP_2024_*of 0.39 (50 citations / 128 articles). In contrast, 20 articles published in 2000 accumulated 1,027 citations over the same period, yielding a *CPP_2024_* of 51 (1,027 citations / 20 articles). As shown in Figure 2, *CPP_2024_* generally stabilizes approximately 16 years after publication. Articles related to dental anxiety show a more prolonged citation impact than those in other dental topics. For instance, studies in dental education ^27^ and temporomandibular disorders ^15^ typically reached their citation plateau within approximately 10 years.

**Figure 2.**
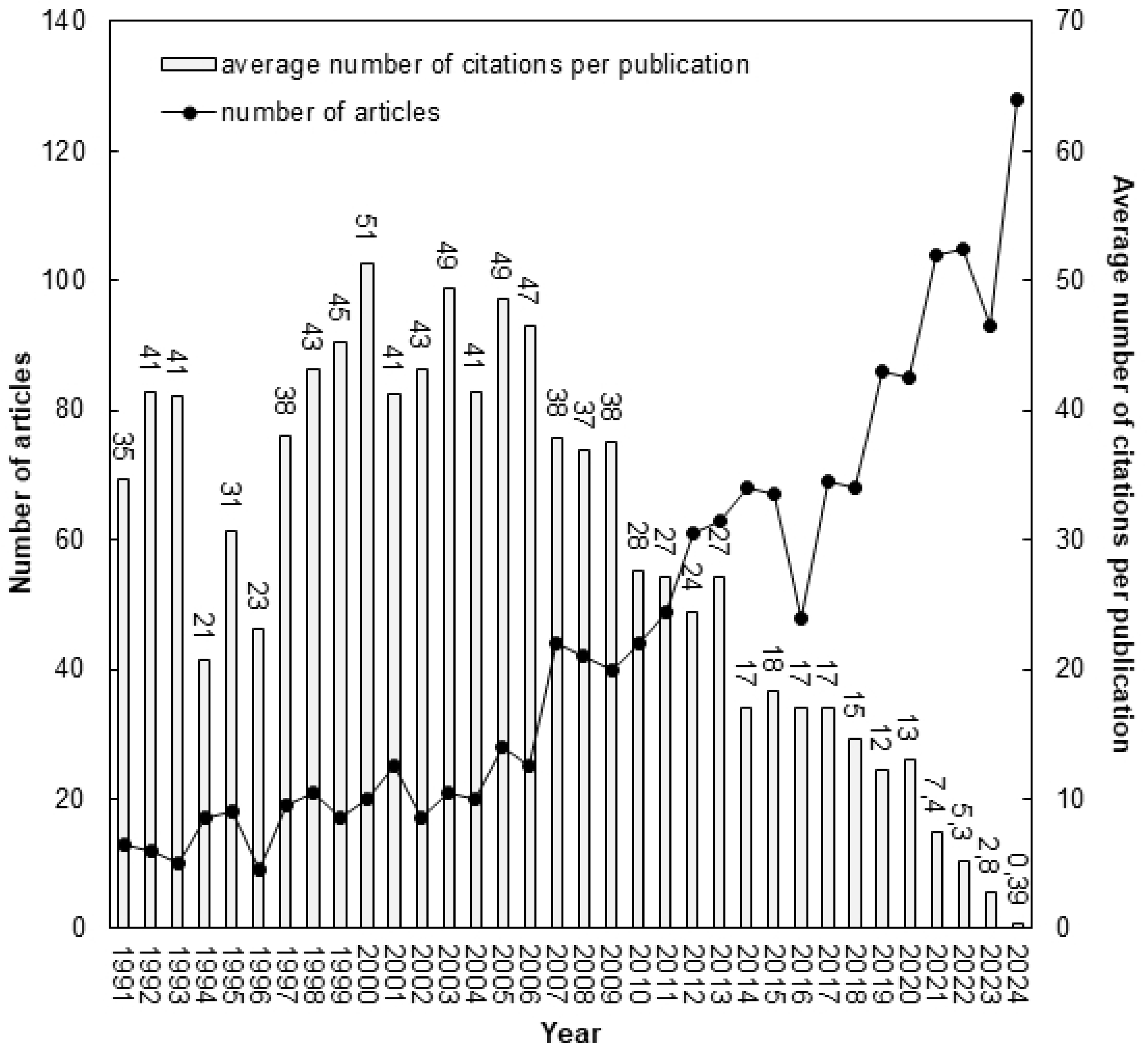
Number of dental anxiety-related articles and average number of citations per publication by year.

A notable increase in the number of publications was observed, rising from 48 articles in 2016 to 128 in 2024. It is likely that this pattern reflects a growing global interest in the topic, with research activity expanding beyond the traditionally dominant countries such as Sweden, Norway, Australia, the Netherlands, the UK, and the USA. Over the past decade, particularly in the last five years, there has been a notable increase in publications from countries such as India, China, and those with Arabic and Persian-speaking populations. While output from the “classical” countries that have historically demonstrated high comparative research output remained relatively stable, the sharp rise in contributions from these emerging regions represents an important and intriguing development.

### Web of Science Category and Journal

In 2023, the Journal Citation Reports (JCR) indexed 9,486 journals within the Science Citation Index Expanded (SCI-EXPANDED), spanning 178 Web of Science categories. Recent studies have established baseline metrics to characterize these categories ^45^ and individual journals, ^46^ using indicators such as the average number of citations per publication (*CPP_year_*) and the average number of authors per publication (*APP*). Within SCI-EXPANDED, a total of 263 journals across 65 Web of Science categories published articles related to dental anxiety. The top 10 most productive categories are summarized in Table 2. Among these, the category *Dentistry, Oral Surgery & Medicine*, which included 91 journals in 2023, accounted for the largest share, publishing 1,147 articles – approximately 74% of the total 1,556 articles on dental anxiety. This substantial proportion reflects the concentrated disciplinary focus of research in this area.

**Table 2.**
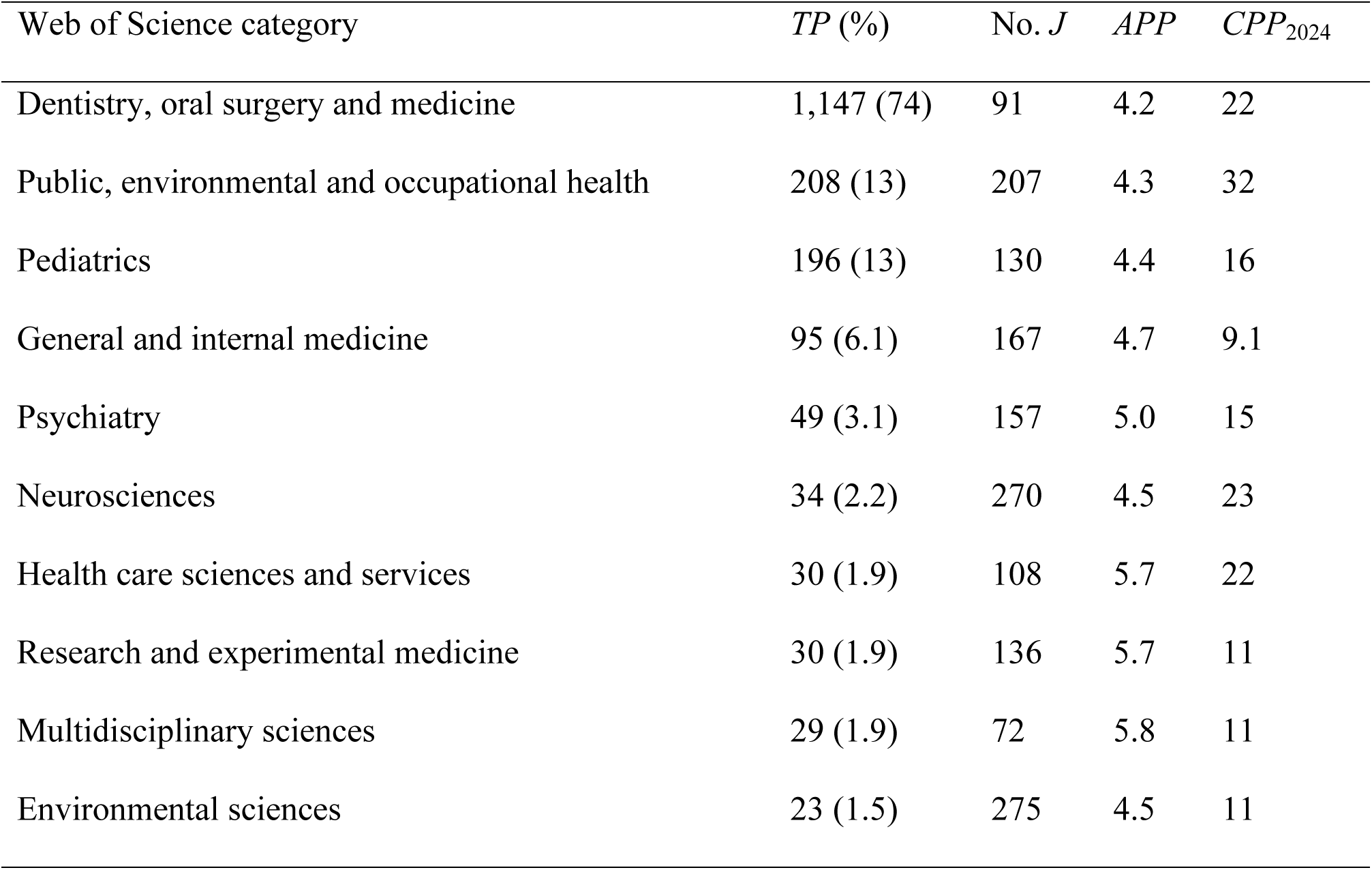
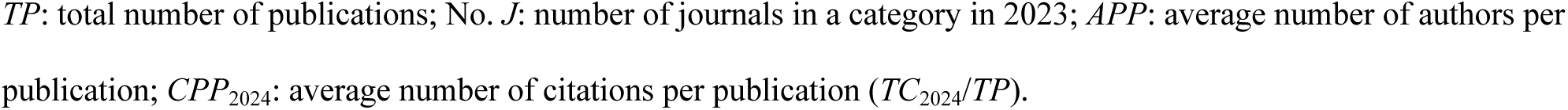
The top 10 most productive Web of Science categories.

| Web of Science category | *TP (%)* | No. *J* | *APP* | *CPP*<sub>2024</sub> |
| --- | --- | --- | --- | --- |
| Dentistry, oral surgery and medicine | 1,147 (74) | 91 | 4.2 | 22 |
| Public, environmental and occupational health | 208 (13) | 207 | 4.3 | 32 |
| Pediatrics | 196 (13) | 130 | 4.4 | 16 |
| General and internal medicine | 95 (6.1) | 167 | 4.7 | 9.1 |
| Psychiatry | 49 (3.1) | 157 | 5.0 | 15 |
| Neurosciences | 34 (2.2) | 270 | 4.5 | 23 |
| Health care sciences and services | 30 (1.9) | 108 | 5.7 | 22 |
| Research and experimental medicine | 30 (1.9) | 136 | 5.7 | 11 |
| Multidisciplinary sciences | 29 (1.9) | 72 | 5.8 | 11 |
| Environmental sciences | 23 (1.5) | 275 | 4.5 | 11 |

$TP$ : total number of publications; No. $J$ : number of journals in a category in 2023; $APP$ : average number of authors per publication; $CPP_{2024}$ : average number of citations per publication ( $TC_{2024}/TP$ ).

Among the top 10 categories listed in Table 2, the category *Public, Environmental & Occupational Health*, comprising 207 journals, achieved the highest *CPP_2024_*, with an average of 32 citations per publication. In contrast, the category *General & Internal Medicine*, with 167 journals, had a *CPP_2024_* of 9.1. In terms of authorship patterns, the category *Multidisciplinary Sciences* exhibited the highest *APP*, with an average of 5.8 authors per publication across 29 articles. Meanwhile, despite its high output, the *Dentistry, Oral Surgery & Medicine* category showed a slightly lower *APP* of 4.2 across 1,147 articles.

Table 3 presents the 11 most productive journals in the field, along with their 2023 impact factor (*IF_2023_*), average number of citations per publication (*CPP_2024_*), and average number of authors per publication (*APP*). All 11 journals are indexed under the *Dentistry, Oral Surgery & Medicine* category. The journal *Community Dentistry and Oral Epidemiology* (*IF_2023_*= 1.8) published the highest number of articles, contributing 123 articles, which accounts for 7.9% of all 1,556 dental anxiety-related articles. This was followed by the journals *European Journal of Oral Sciences* and *BMC Oral Health*.

**Table 3.**
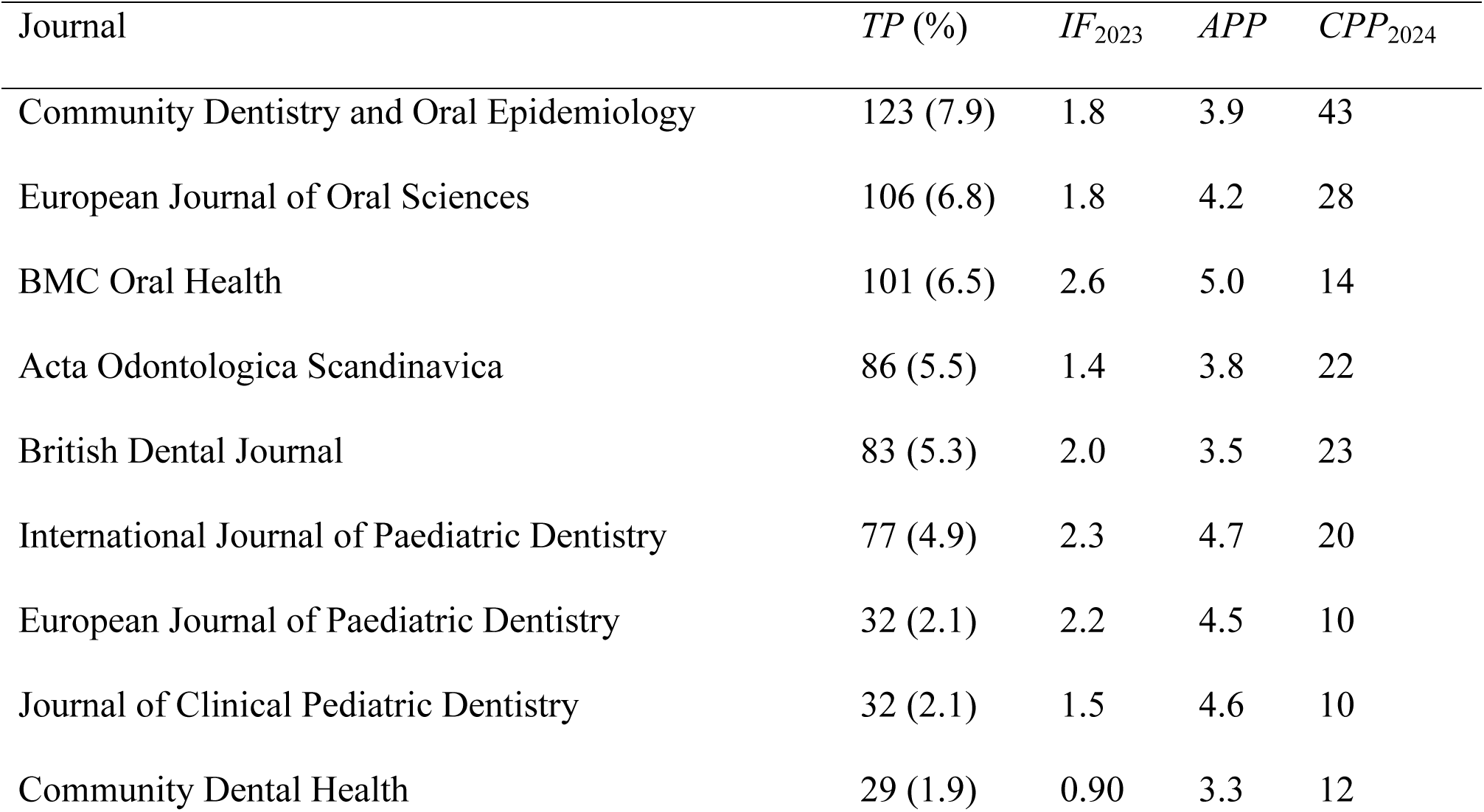
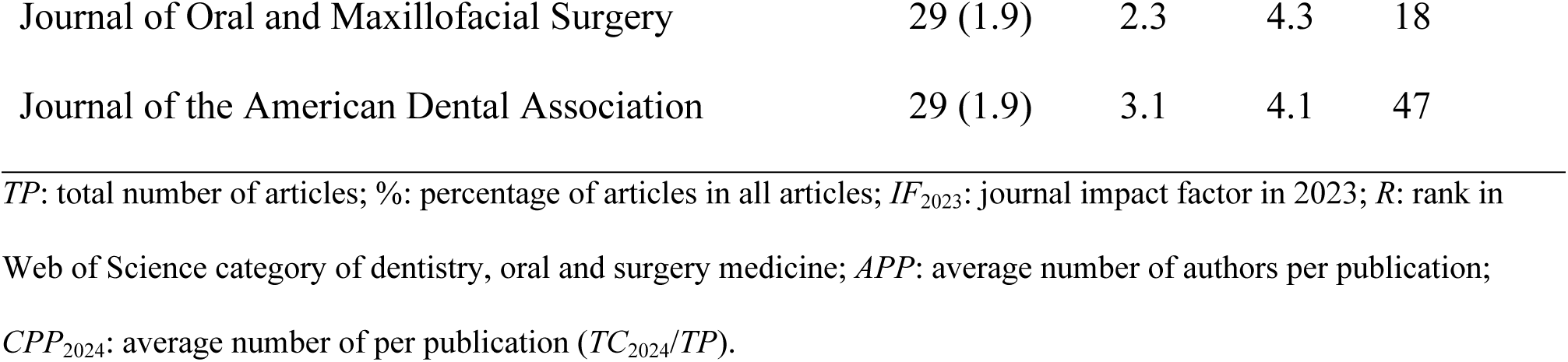
The top 11 most productive journals.

Among the top journals, *the Journal of the American Dental Association* recorded the highest *CPP_2024_*, with an average of 47 citations per article. In contrast, *the European Journal of Paediatric Dentistry* (*IF_2023_* = 2.2) published 32 articles, with a comparatively lower *CPP_2024_* of 10. *APP* values among the top 11 journals ranged from 5.0 authors per article in the journal *BMC Oral Health* to 3.3 in the journal *Community Dental Health*.

Outside the top 11 in terms of productivity, several journals with limited contributions exhibited notably high impact factors. The journal *Advanced Functional Materials* had the highest *IF_2023_* of 19, based on a single article. This was followed by the journals *Psychotherapy and Psychosomatics* (two articles; *IF_2023_* = 16) and *the International Journal of Surgery* (one article; *IF_2023_* = 13).

### Publication performances: countries and institutions

It is widely recognized that the first and corresponding authors typically contribute most substantially to a research article. ^47^ At the institutional level, the affiliation of the corresponding author is often regarded as indicative of the study’s origin or home institution. ^18^ Within the SCI-EXPANDED database, six dental anxiety-related articles (0.39% of the 1,556 articles) lacked affiliation data. The remaining 1,550 articles were authored by researchers affiliated with institutions in 79 countries. Of these, 1,261 articles (81%) were classified as single-country publications, originating from 60 countries, and had an average of 20 citations per publication (*CPP_2024_*). The other 289 articles (19%) were produced through international collaborations involving authors from 69 countries, and these had a slightly higher *CPP_2024_* of 21. These findings suggest that international collaboration may contribute modestly to increased citation impact in dental anxiety research.

Six publication indicators, along with their corresponding citation indicators (*CPP_2024_*), were used to compare the top 10 most productive countries in dental anxiety research, following the framework proposed by Ho and Mukul in 2021. ^38^ These countries include five from Europe, two from the Americas, two from Asia, and one from Oceania (Table 4). Egypt, with 26 articles (ranked 20^th^ overall), was the most productive country from Africa.

**Table 4.**
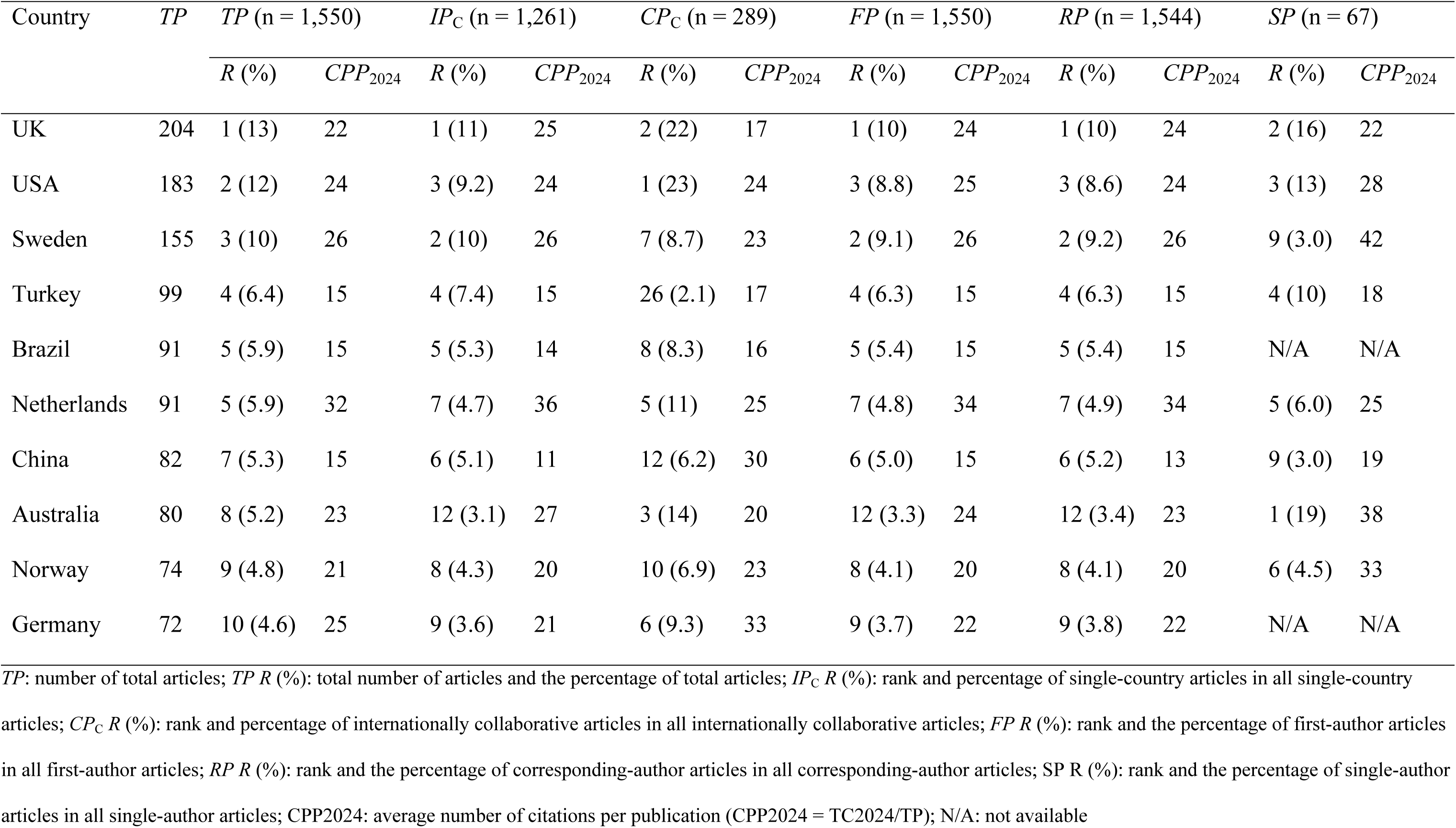
Top 10 productive countries.

The United Kingdom led in four of the six publication indicators: total publications (*TP* = 204; 13% of the 1,550 articles), single-country publications (*IP_C_* = 139; 11% of the 1,261 single-country articles), first-author publications (*FP* = 156; 10% of the 1,550 first-author articles), and corresponding-author publications (*RP* = 162; 10% of the 1,544 corresponding-author articles). The United States ranked highest in internationally collaborative publications (*CP_C_*), contributing 67 articles, or 23% of the 289 collaborative articles. Australia led in single-author publications (*SP*), with 13 articles, accounting for 19% of the 67 single-author articles.

Among the top 10 most productive countries listed in Table 4, the Netherlands exhibited the highest citation impact across four key publication indicators. With 91 total publications (*TP*), 59 single-country publications (*IP_C_*), 75 first-author publications (*FP*), and 75 corresponding-author publications (*RP*), the Netherlands achieved the highest *CPP_2024_* values of 32, 36, 34, and 34 citations per publication, respectively. Germany led in internationally collaborative publications (*CP_C_*), with 27 such articles achieving an average of 33 citations per publication—the highest in this category. Sweden, despite contributing only two single-author articles (*SP*), recorded the highest *CPP_2024_* for that indicator, with 42 citations per publication.

Of the 1,550 analyzed dental anxiety-related articles, 562 (36%) were single-institution publications, with a *CPP_2024_* of 21 citations per article, while the remaining 988 articles (64%) resulted from institutional collaborations, with a slightly lower *CPP_2024_* of 20. This suggests that institutional collaboration did not enhance—and may have slightly reduced—citation impact in this research area.

Table 5 presents the top 10 most productive institutions and their publication profiles. Among these, institutions from the Netherlands, Norway, and Sweden were each represented twice, while Australia, Finland, the United Kingdom, and the United States were each represented once.

**Table 5.**
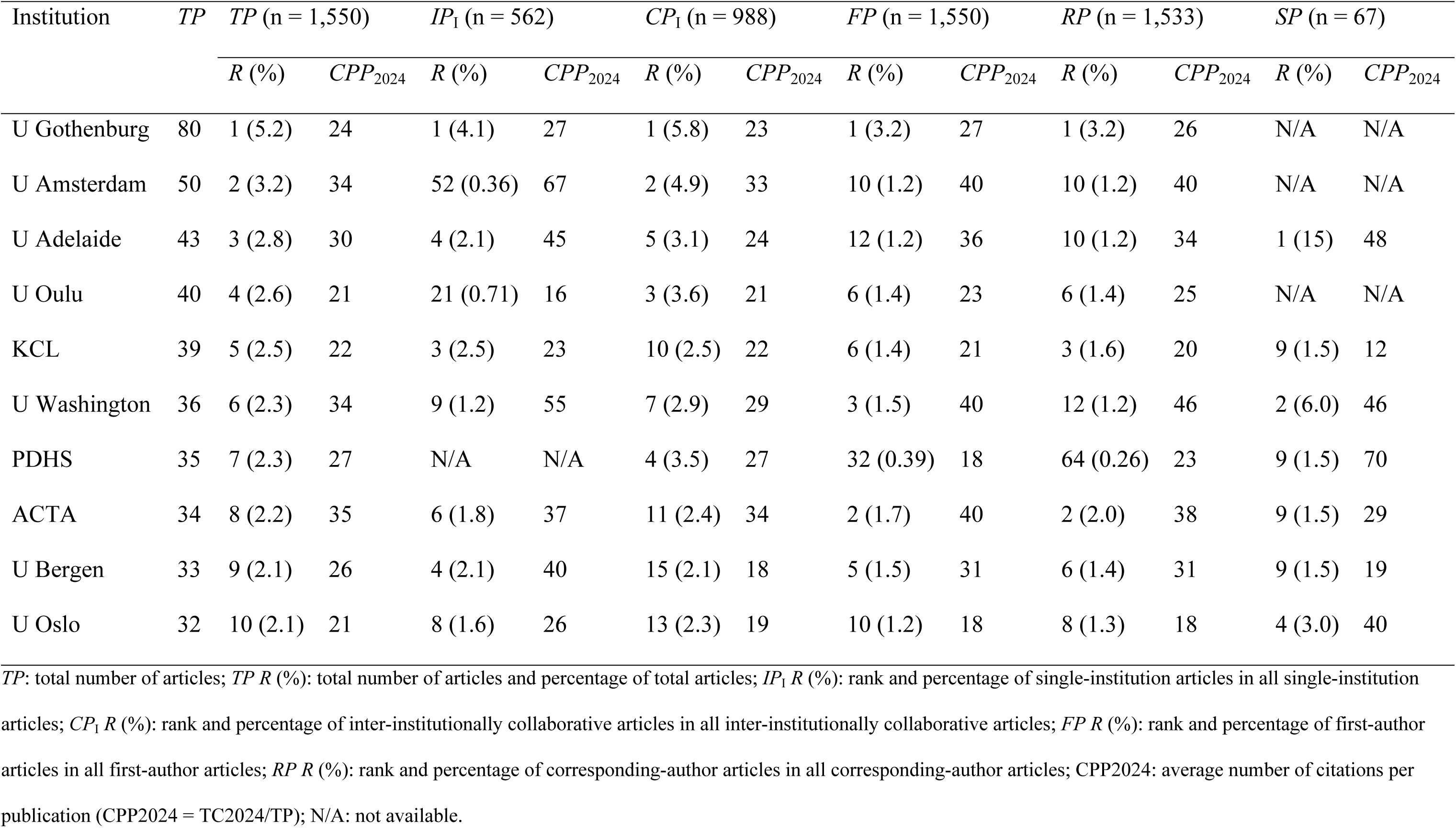
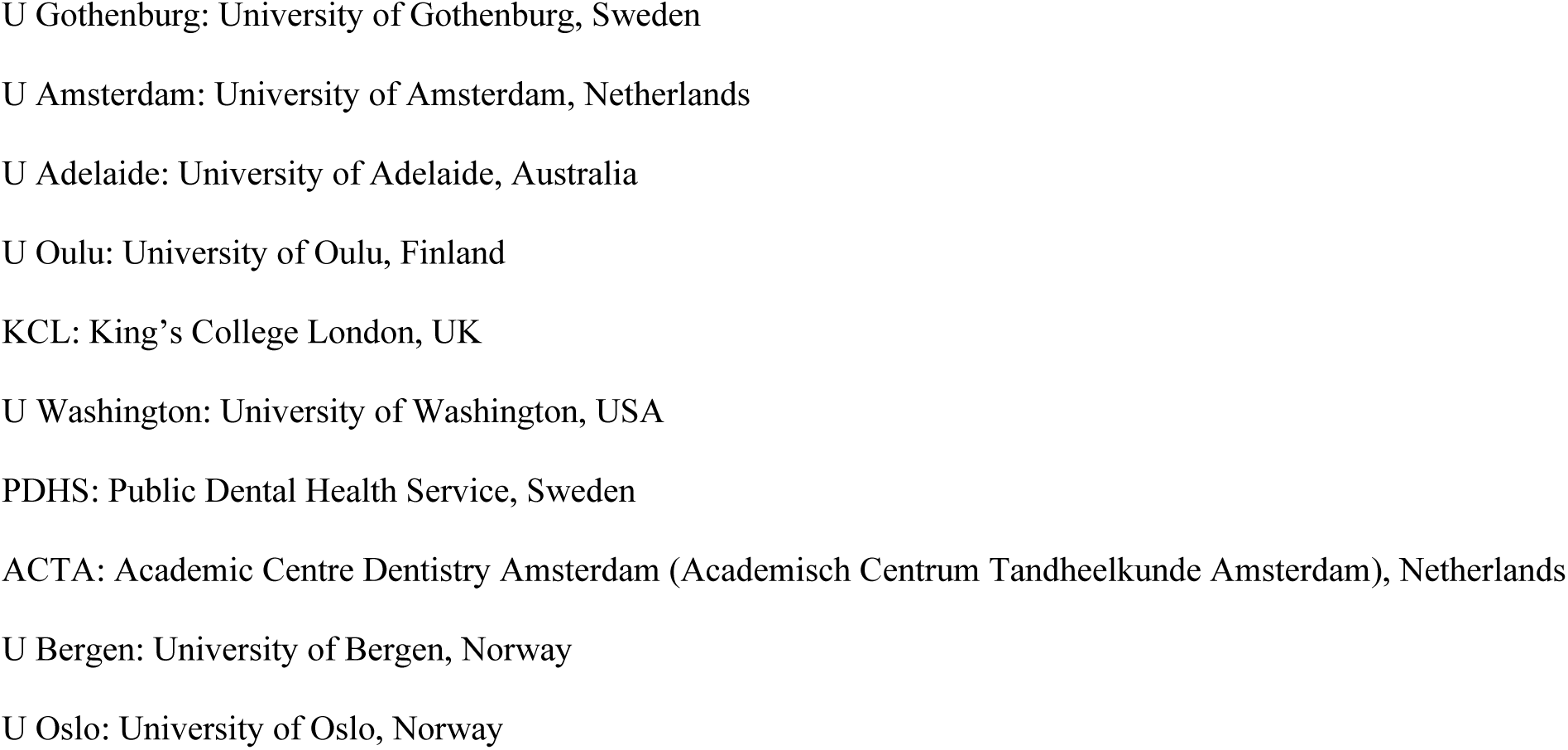
Top 10 most productive institutions.

Notably, three institutions – the University of Gothenburg (Sweden), the University of Amsterdam (Netherlands), and the University of Oulu (Finland) – produced single-author articles.

The University of Gothenburg (U Gothenburg) led in five of the six publication indicators: total publications (*TP* = 80; 5.2% of the 1,550 articles), single-institution publications (*IP_I_* = 23; 4.1% of the 562 single-institution articles), inter-institutional collaborations (*CP_I_* = 57; 5.8% of the 988 collaborative articles), first-author publications (*FP* = 49; 3.2% of the 1,550 first-author articles), and corresponding-author publications (*RP* = 49; 3.2% of the 1,533 corresponding-author articles). The University of Adelaide (U Adelaide) led in single-author publications (*SP*), contributing 10 articles, or 15% of the 67 single-author articles.

Among the top 10 institutions, the Academic Centre for Dentistry Amsterdam (ACTA) had the highest *CPP_2024_* for both total publications (*TP* = 34; *CPP_2024_* = 35) and inter-institutional collaborations (*CP_I_* = 24; *CPP_2024_* = 34). The University of Washington (U Washington) recorded the highest citation impact for first-author and corresponding-author publications, with *CPP_2024_* values of 40 (*FP* = 24) and 46 (*RP* = 18), respectively. The University of Amsterdam (U Amsterdam) had the highest *CPP2024* for collaborative publications (*CP_I_* = 48; *CPP2024* = 67), while the Public Dental Health Service in Sweden (PDHS) led in citation impact for single-author publications, with one article cited 70 times.

These findings are not unexpected, as it is well established that the United States and United Kingdom – along with, to a lesser extent, Australia and the Netherlands – are recognized leaders in psychology and psychiatry research, ^48^ subjects that converge with dental anxiety. Notably, is that the Nordic countries, despite their relatively small populations, have a disproportionately high impact on dental anxiety research. This is largely attributable to certain specific impactful institutions and individuals who have built strong research traditions in the field.

### Citation histories of the ten most frequently cited articles

Total citation counts in the Web of Science Core Collection (WoSCC) are continually updated. To minimize potential bias associated with the dynamic nature of citation data, the total number of citations from the year of publication through the most recently completed year (*TC_year_*) was used, as recommended by Wang et al. in 2011, ^34^ to ensure a more comprehensive and consistent dataset. The citation trajectories of the ten most highly cited articles on dental anxiety are illustrated in Figures 3 and 4. It is important to note that highly cited articles do not always maintain high annual citation rates over time. ^41^ For instance, the article titled “Are there differences in oral health and oral health behavior between individuals with high and low dental fear” by Schuller et al. (2003) accumulated a total of 161 citations by 2024 (*TC_2024_*), ^49^ ranking 9th among dental anxiety-related articles. However, in 2024, it received only five citations (*C_2024_*), placing it at 174th for that year (Fig. 4). Similarly, the article by Milgrom et al. (1997) also demonstrated low recent-year impact, ^50^ with a *C_2024_* of six citations.

**Figure 3.**
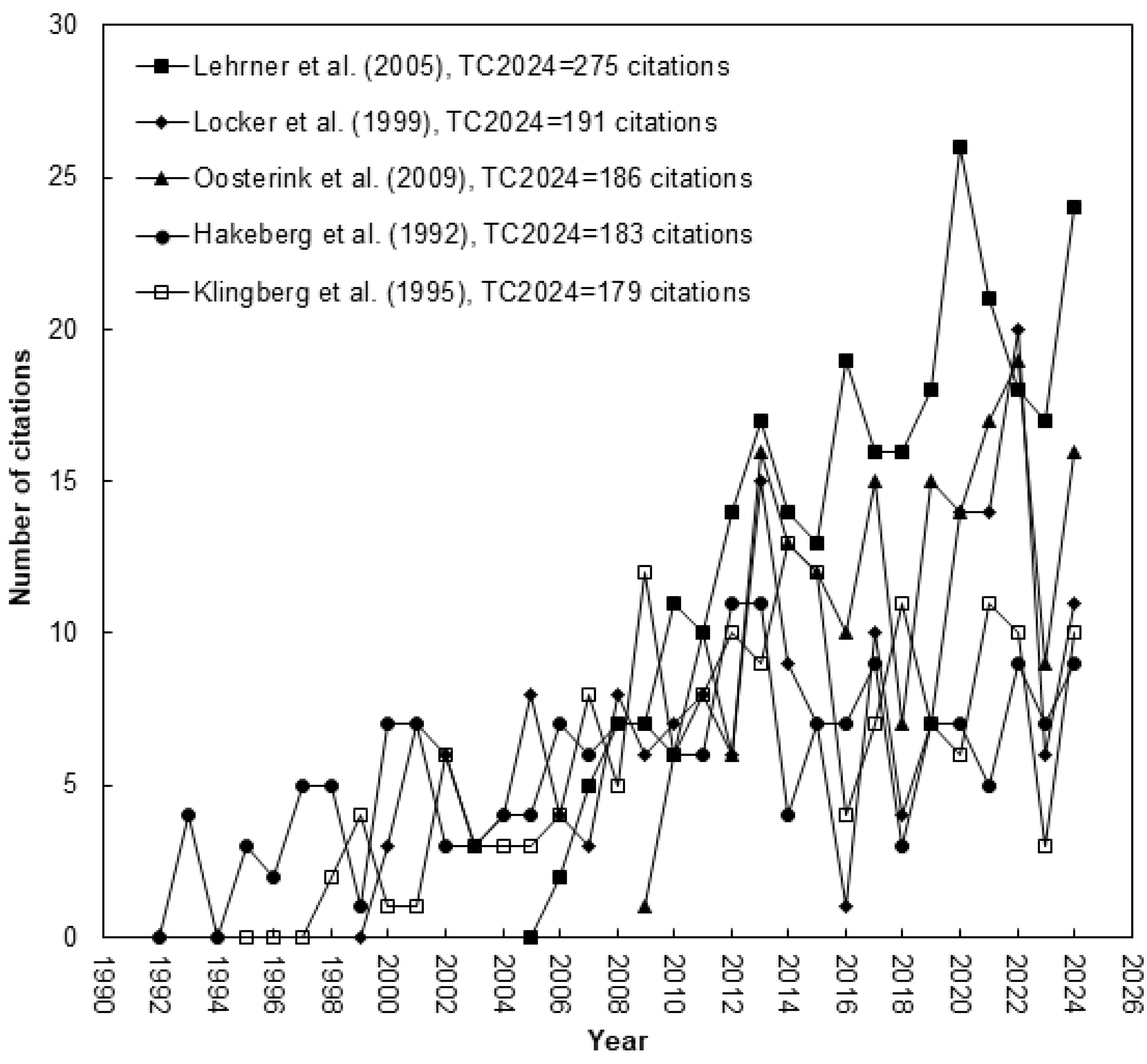
Citation histories of the top five most frequently cited articles.

**Figure 4.**
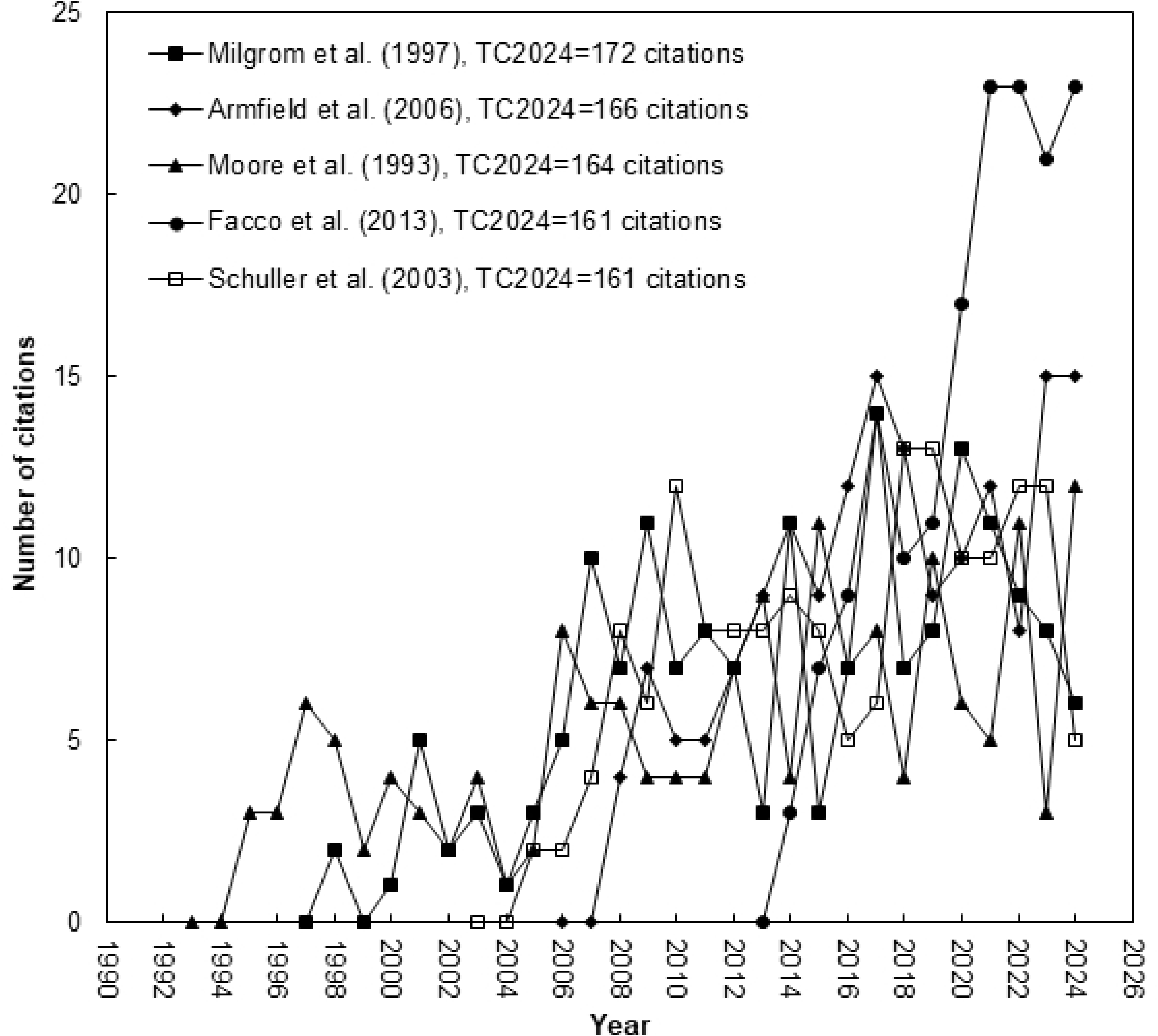
Citation histories of the top 6-10 most frequently cited articles.

Table 6 lists the ten most frequently cited articles. The citation indicator *TC_2024_*was employed to identify the most highly cited articles overall, while *C_2024_*was used to assess the articles with the highest citation impact in the most recent year within the field. Notably, only four of the ten most frequently cited articles also appeared among the top ten most impactful articles in 2024. These four articles are summarized below:

**Table 6.** Top ten most frequently cited dental anxiety-related articles.

| Rank<br>($TC_{2024}$) | Rank<br>($C_{2024}$) | Title | Country | Reference |
| --- | --- | --- | --- | --- |
| 1 (275) | 1 (24) | Ambient odors of orange and lavender reduce anxiety and improve mood in a dental office | Austria, Germany | Lehrner et al. (2005) |
| 2 (191) | 28 (11) | Age of onset of dental anxiety | Canada | Locker et al. (1999) |
| 3 (186) | 5 (16) | Prevalence of dental fear and phobia relative to other fear and phobia subtypes | Netherlands | Oosterink et al. (2009) |
| 4 (183) | 52 (9) | Prevalence of dental anxiety in an adult-population in a major urban area in Sweden | Sweden | Hakeberg et al. (1992) |
| 5 (179) | 41 (10) | Child dental fear: Cause-related factors and clinical effects | Sweden | Klingberg et al. (1995) |
| 6 (172) | 128 (6) | Four dimensions of fear of dental injections | USA | Milgrom et al. (1997) |
| 7 (166) | 9 (15) | Dental fear in Australia: who's afraid of the dentist | Australia | Armfield et al. (2006) |
| 8 (164) | 19 (12) | Prevalence and characteristics of dental anxiety in Danish adults | China, Denmark | Moore et al. (1993) |
| 9 (161) | 174 (5) | Are there differences in oral health and oral health behavior between individuals with high and low dental fear | Netherlands,<br>Norway | Schuller et al. (2003) |
| 9 (161) | 2 (23) | Validation of visual analogue scale for anxiety (VAS-A) in preanesthesia evaluation | Italy | Facco et al. (2013) |

$TC_{2024}$ : total number of citations from WoSCC since publication year to the end of 2024; $C_{2024}$ : number of citations of an article in 2024 only.

1. *Ambient odors of orange and lavender reduce anxiety and improve mood in a dental office* ^51^

The article published by five authors from Austria and Germany with a *TC*_2024_ of 275 citations (rank 1^st^) and a *C*_2024_ of 24 citations (rank 1^st^). This study investigated whether ambient odors of orange and lavender could reduce anxiety and improve mood among patients in a dental office setting – a context frequently associated with elevated stress. Building on previous research suggesting that certain essential oils may influence emotional states, the authors designed a controlled, between-subjects experiment involving 200 dental patients aged 18 to 77 years. Participants were assigned to one of four groups: exposure to orange odor, lavender odor, uplifting music, or no sensory stimulation (control). While waiting for dental treatment, patients completed validated self-report measures assessing state anxiety and emotional well-being, including mood, alertness, and calmness. Statistical analyses revealed that both the orange and lavender odor conditions led to significantly lower levels of state anxiety and higher ratings of mood and calmness compared to the control group. In contrast, music did not significantly differ from the control condition on these measures. No significant group differences were found for alertness, and no interactions with gender were detected. These findings provide empirical support for the sedative and mood-enhancing properties of essential oils before a dental visit and suggest that incorporating natural ambient odors in dental offices may offer a simple, non-invasive method to reduce anticipatory anxiety and promote emotional well-being in a non-clinical population before a dental visit.

2. *Prevalence of dental fear and phobia relative to other fear and phobia subtypes* ^52^

The article published by three authors from Netherlands with a *TC*_2024_ of 186 citations (rank 3^rd^) and a *C*_2024_ of 16 citations (rank 5^th^). This study is among the earliest to examine dental phobia - a distinct medical condition - rather than dental anxiety, within a large and representative population sample. The study assessed the point prevalence of dental fear and dental phobia in comparison to ten other common fears and subtypes of specific phobia, based on DSM-IV-TR diagnostic criteria.

A representative sample of 1,959 Dutch adults (aged 18–93) completed a structured questionnaire evaluating the presence, severity, and psychological correlates of various fears. Participants who endorsed a specific fear completed a validated Phobia Checklist assessing DSM-based criteria and trauma-related symptoms, including intrusive re-experiencing. The results showed that 24.3% of participants reported dental fear, making it the fourth most common fear after snakes, heights, and physical injuries. Dental phobia was identified in 3.7% of the sample, making it the most prevalent specific phobia subtype. Dental fear was also rated as the most severe and was most strongly associated with intrusive re-experiencing (49.4%), suggesting a potential trauma-related dimension. Women more frequently reported fear of dental treatment, although gender differences were not significant for dental phobia prevalence. The findings underscore that while dental fear, especially the sever form of dental phobia, is not the most widespread fear, it appears to be among the most intense and psychologically burdensome. The authors highlight the need to recognize dental phobia as a serious and distinct mental health condition that may warrant specialized clinical attention

3. *Dental fear in Australia: who’s afraid of the dentist*^53^

The article published by three authors from the University of Adelaide in Australia with a *TC*_2024_ of 166 citations (rank 7^th^) and a *C*_2024_ of 15 citations (rank 9^th^). The purpose of this study was to determine the prevalence of dental fear in Australia and to explore its association with demographic, socio-economic, oral health, insurance, and dental service utilization factors. Using data from the 2002 National Dental Telephone Interview Survey, researchers conducted structured telephone interviews with a nationally representative sample of 7,312 individuals aged five years and older. Dental fear was assessed using a validated single-item question. Results indicated that 16.1% of the Australian population reported high dental fear, with higher rates observed among women, individuals aged 40–64, and those from lower socio-economic backgrounds. Dentate individuals reported significantly more fear than those who were edentulous, and people with high dental fear tended to have more missing teeth, less frequent dental visits, and longer intervals since their last dental appointment. Interestingly, while dental insurance coverage was only slightly lower among individuals with high fear, time since last dental visit showed a strong association, with fear increasing as time since the last visit lengthened. The study also found that people with lower income, lower education, and unstable employment were more likely to report high dental fear.

These authors concluded that the findings underscore the need for tailored public health strategies and clinical interventions aimed at identifying and supporting individuals at greater risk of dental fear and avoidance.

4. *Validation of visual analogue scale for anxiety (VAS-A) in preanesthesia evaluation*^54^

The article published by seven authors from the University of Padua in Italy with a *TC*_2024_ of 161 citations (rank 9^th^) and a *C*_2024_ of 23 citations (rank 2^nd^). In this validated the Visual Analogue Scale for Anxiety (VAS-A) as a simple and reliable tool for assessing preoperative anxiety in patients undergoing oral surgery. While VAS-A has been used in several clinical contexts, its accuracy and optimal cutoff threshold had not been clearly established. One hundred patients (median age 49) scheduled for implantology, or dental extractions completed the VAS-A alongside the Corah Dental Anxiety Scale (CDAS), the State-Trait Anxiety Inventory (STAI-Y1 and Y2), and the Beck Depression Inventory (BDI). The study found significant correlations between VAS-A and CDAS, STAI-Y1, and STAI-Y2, although data dispersion was noted. VAS-A did not significantly correlate with BDI, but it identified 5 out of 7 participants with depressive symptoms. ROC curve analysis suggested 46 mm as the optimal threshold for clinically relevant anxiety, showing high sensitivity (83%) and good negative predictive value (87%), though with lower specificity (61%).

Interestingly, VAS-A tended to overestimate anxiety compared to other tools, which the authors suggest may reflect its ability to capture broader anxiety components, including fear of surgical outcomes. The findings support the use of VAS-A as a fast, practical, and sensitive screening tool for preoperative anxiety, making it particularly valuable in clinical anesthetic settings with limited time for psychological assessment

The four articles with the most impact during the last year of publishing had only a few annual citations during their first years of publication. Notably, the citation trajectories began to rise significantly around 2010, regardless of their original publication dates. This trend aligns with the broad surge of academic interest in dental anxiety, as evidenced by the growing volume of literature on the topic in recent years. The studies by Oostering et al. (2009) and Armfield et al. (2006) have been particularly impactful in establishing the epidemiological and psychological underpinnings of dental anxiety and dental phobia. ^52,53^ Their work on prevalence, correlations, and psychosocial consequences has sustained long-term relevance and continues to inform research. In contrast, the two more recent contributions by Lehrner et al. (2005) and Facco et al. (2013) have introduced innovative methods that reflect emerging trends in the field. ^51,54^ Facco et al. (2013) described an accessible method for quantifying dental anxiety in clinical research. ^54^ Lehrner et al. (2005) were among the first to empirically demonstrate the potential of aromatherapy as a non-invasive intervention for reducing dental anxiety. ^51^ These studies were at the forefront of a notable shift toward intervention-focused research, and a marked increase in publications from countries such as India and China, where interest in complementary approaches, such as aromatherapy and other comfort-enhancing modalities, has grown substantially. Taken together, these four papers represent articles on different important topics of dental anxiety and their foundational role in shaping contemporary research directions, methodological approaches, and clinical practices.

### Research foci

Over the past 15 years, Ho’s research group has developed innovative methodologies for analyzing word distributions in article titles, abstracts, author keywords, and Keywords Plus to identify research foci and emerging trends. ^21,55^ The extracted keywords were subsequently compiled into a word bank, which formed the basis for identifying major thematic areas and tracing their evolution over time. ^55^ The 1,556 articles included in this study were distributed across three distinct periods, each comprising a comparable number of publications: 511 articles from 1991 to 2011, 530 articles from 2012 to 2019, and 515 articles from 2020 to 2024. Excluding the original search terms, the 21 most frequently used author keywords in dental anxiety research and their distribution across the three sub-periods (1991–2011, 2012–2019, and 2020–2024) are presented in Table 7.

**Table 7.** Top 21 most frequency used author keywords during 1991-2024.

| Author keywords | *TP* | 91-24 *R* (%) | 91-11 *R* (%) | 12-19 *R* (%) | 20-24 *R* (%) |
| --- | --- | --- | --- | --- | --- |
|  |  | n = 1,556 | n = 511 | n = 530 | n = 515 |
| Oral health | 105 | 1 (8.3) | 2 (5.7) | 1 (7.3) | 1 (11) |
| Children | 84 | 2 (6.6) | 5 (4.4) | 1 (7.3) | 2 (7.8) |
| Pain | 75 | 3 (5.9) | 2 (5.7) | 3 (5.9) | 3 (6.1) |
| Dental care | 56 | 4 (4.4) | 1 (7.3) | 7 (3.8) | 10 (2.6) |
| Dental caries | 51 | 5 (4.0) | 7 (3.4) | 6 (4.2) | 5 (4.4) |
| Epidemiology | 43 | 6 (3.4) | 4 (5.2) | 5 (4.5) | 47 (0.87) |
| Child | 42 | 7 (3.3) | 18 (1.8) | 4 (4.7) | 8 (3.3) |
| Pediatric dentistry | 38 | 8 (3.0) | 37 (1.0) | 8 (3.1) | 4 (4.6) |
| Quality of life | 29 | 9 (2.3) | 18 (1.8) | 10 (2.3) | 10 (2.6) |
| Adolescents | 26 | 10 (2.1) | 12 (2.1) | 10 (2.3) | 19 (1.7) |
| Prevalence | 25 | 11 (2.0) | 9 (2.9) | 17 (1.9) | 29 (1.3) |
| Depression | 24 | 12 (1.9) | 37 (1.0) | 9 (2.6) | 13 (2.0) |
| Sedation | 24 | 12 (1.9) | 28 (1.3) | 26 (1.4) | 9 (2.8) |
| Oral Health-related quality of life | 23 | 14 (1.8) | 37 (1.0) | 10 (2.3) | 13 (2.0) |
| Psychometrics | 23 | 14 (1.8) | 6 (4.2) | 19 (1.6) | N/A |
| Virtual reality | 23 | 14 (1.8) | 169 (0.26) | 75 (0.7) | 6 (4.1) |
| Dental treatment | 22 | 17 (1.7) | 28 (1.3) | 13 (2.1) | 19 (1.7) |
| Behavioral science | 21 | 18 (1.7) | 7 (3.4) | 19 (1.6) | 248 (0.22) |
| Caries | 20 | 19 (1.6) | 18 (1.8) | 49 (0.94) | 13 (2.0) |
| Psychology | 20 | 19 (1.6) | 10 (2.6) | 26 (1.4) | 47 (0.87) |
| Reliability | 20 | 19 (1.6) | 12 (2.1) | 13 (2.1) | 80 (0.65) |

*TP*: number of articles; %: percentage in each period; *R*: rank in each period.

The authors identified five overarching subjects in the research of dental anxiety: (1) Etiology and Risk Factors, (2) Clinical Presentation and Consequences of Dental Anxiety, (3) Prevalence and Assessment Tools, (4) Treatment and Preventive Interventions, (5) Pediatric dentistry.

*(1) Etiology and Risk Factors* deal with the origins and causes of dental anxiety. The topic investigates how dental anxiety develops through a multifactorial interplay of psychological, environmental, and biological factors. Key contributors include negative past dental experiences, trait anxiety, and other personal factors, and forms of vicarious and observational learning. Identifying these risk factors is critical for understanding why dental anxiety develops and can therefore inform the design of preventive interventions aimed at reducing its incidence.

**Supporting words:** Disorders, Experience, Acquisition, Experiences, Onset, Predictors, Origins, Memory, Childhood, Conditioning experiences, Personality, Etiology, Cognitive vulnerability, Determinants, Risk-factors

*(2) Clinical presentation and consequences of dental anxiety* cover how dental anxiety manifests itself and its consequences. The clinical presentation is characterized by heightened autonomic arousal, avoidance behaviors, and emotional distress. Many times, it is also accompanied by cognitive patterns such as catastrophizing, attention biases, and negative appraisals. The avoidance behaviors lead to delayed or avoided dental visits, resulting in the progression of oral diseases, increased treatment complexity, and a diminished quality of life. Furthermore, this cycle of avoidance maintains or reinforces the anxiety, creating a self-perpetuating pattern that poses substantial challenges for both patients and practitioners.

**Supporting words:** Avoidance, Health, Impact, Oral health, Consequences, Oral health-related quality of life, Symptoms, Responses, Cognitions

*(3) Prevalence and Assessment Tools* investigate how widespread dental anxiety is in a population, and the development of instruments for accurate identification. Epidemiological studies consistently report high prevalence rates, with variations influenced by age, gender, and cultural context. The development and validation of psychometric instruments aids in this, but the instruments measuring dental anxiety are also used as outcome measurements in intervention studies.

**Supporting words:** Prevalence, Scale, Validity, Reliability, Population, Validation, Version, Psychometrics, Subscale, Questionnaire, Survey schedule, Anxiety scale, Fear survey schedule, Psychometric properties, Adaptation, Fear survey, Dental anxiety scale

*(4) Treatment and Preventive Interventions* evaluate how dental anxiety can be treated or prevented in different populations. The treatment and prevention of dental anxiety encompass a diverse range of interventions aimed at reducing anxiety levels and improving patient engagement with dental care. These strategies include psychological approaches such as exposure-based cognitive-behavioral therapy (CBT) for more severe dental anxiety and dental phobia, dental behavioral support (management), as well as pharmacological options like sedation and anxiolytics. Additionally, non-traditional modalities, such as aromatherapy and virtual reality, have gained increasing attention as adjunctive measures in recent years. Preventive efforts focus on early identification, patient education, and the implementation of anxiety-reducing practices within dental environments.

**Supporting words:** Management, Sedation, Virtual reality, Therapy, Midazolam, Efficacy, Anesthesia, Sedation, Intervention, Follow-up, Nitrous-oxide, Premedication, Prevention, Relaxation, Strategies, Propofol, Acupuncture, Applied relaxation, Audiovisual distraction, Virtual-reality, Conscious sedation, Randomized controlled-trial, Applied tension, Aromatherapy, Cognitive therapy, Group-therapy, Individual desensitization, Nitrous-oxide sedation, Cognitive-behavioral therapy, Controlled-trial, Dexmedetomidine, Diazepam, Double-blind, Hypnosis, Interventions, Lavender

*(5) Pediatric dentistry* focuses on dental anxiety in children and adolescents. Pediatric dentistry is the dental specialty that has shown sustained interest in the study and management of dental anxiety, as this condition frequently originates in childhood and demonstrates its highest prevalence among pediatric populations. Addressing dental anxiety is an integral component of pediatric dental care, and specialists in pediatric dentistry often treat patients unable to receive dental care from the general practitioners. Early dental experiences are critical in shaping long-term attitudes toward oral health, with negative encounters often serving as precursors to persistent dental fear and avoidance behaviors. This intersection has positioned dental anxiety as a key focus of research within the specialty, prompting continued research into age-appropriate assessment tools, intervention, and preventive strategies tailored to children and adolescents.

**Supporting words:** Children, Child, Pediatric dentistry, Adolescents, Behavior management problems, Management problems, Distraction, Pediatric dentistry

The development trends of the five primary research topics in dental anxiety, as presented in Figure 5, illustrate notable shifts and progressions within the field over the past three decades, highlighting implications for future research and clinical practice.

**Figure 5.**
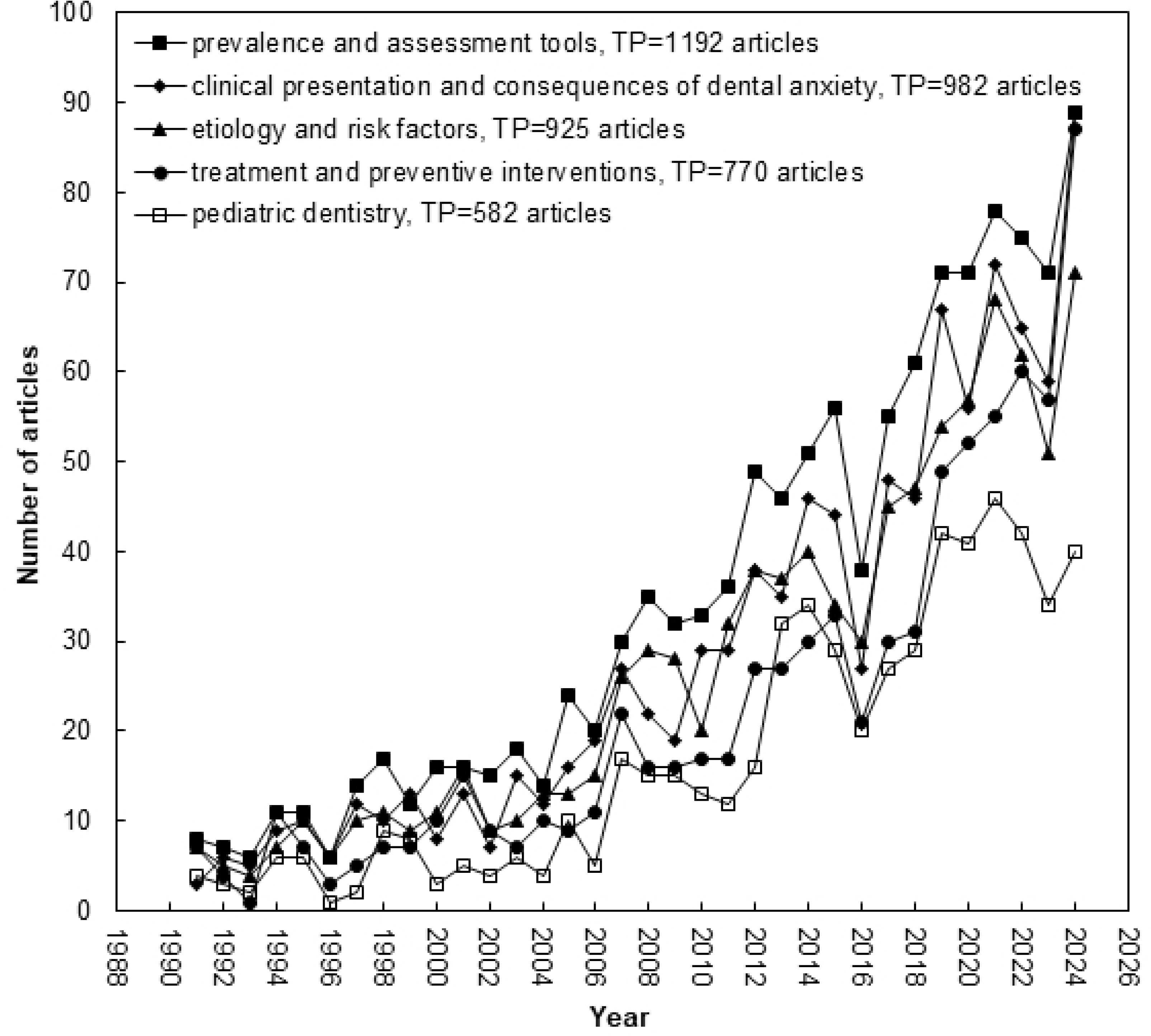
Development trends for five main research topics in dental anxiety research.

The first topic, *Etiology and Risk Factors*, has experienced sustained scholarly attention throughout the periods examined, reflecting the fundamental importance of understanding the origins of dental anxiety. Research consistently underscores the multifactorial nature of dental anxiety, emphasizing psychological variables such as past traumatic dental experiences and cognitive vulnerabilities, including catastrophic thinking and memory biases. ^8,56^ Recent literature suggests an increased focus on cognitive and conditioning experiences in childhood, highlighting the critical role of early experiences in shaping long-term dental anxiety. ^4^ This underscores the importance of preventive strategies initiated early in life to mitigate future anxiety.

In terms of *Clinical Presentation and Consequences of Dental Anxiety*, the research has progressively expanded, reflecting increased recognition of how dental anxiety negatively impacts both oral health and overall quality of life. Avoidance behaviors, commonly associated with dental anxiety, have been shown to effect anxiety and exacerbate oral health deterioration, thus compounding the complexity and invasiveness of required treatments. ^57,58^

The third topic, *Prevalence and Assessment Tools*, remains critically important, with consistent advancements in methodological accuracy and instrument development. Studies show variations in prevalence based on demographic and cultural factors, and although the reliability of psychometric tools is generally high, the construct of dental anxiety that the instruments measure varies significantly. ^56,59^ The continual refinement of assessment instruments enhances early detection and accurate measurement of dental anxiety, fundamental for both epidemiological research and intervention effectiveness.

Research related to *Treatment and Preventive Interventions* has exhibited a marked increase, particularly from 2012 and on. This likely reflects greater integration of multidisciplinary approaches, combining psychological, pharmacological, and novel adjunctive therapies such as virtual reality and aromatherapy. ^60,61^ The rise of cognitive-behavioral therapy strategies and the use of different sedation methods highlights a shift towards individualized treatment plans to enhance patient comfort and treatment outcomes.

Finally, the field of *Pediatric Dentistry* demonstrates significant and consistent growth in interest, reflecting increased recognition of childhood as a critical period for addressing dental anxiety.

Pediatric dentistry has prioritized behavioral management strategies and preventive interventions aimed at reducing anxiety and improving children’s dental experiences, as negative early encounters can predispose individuals to chronic dental fear. ^5^ Future research will likely continue emphasizing tailored interventions that accommodate developmental stages and specific clinical groups such as those with neurodevelopmental disorders, thereby promoting long-term positive attitudes toward dental care.

Taken together, the historical trends observed in Figure 6 emphasize a clear progression towards a more integrated, patient-centered, and multidisciplinary approach to dental anxiety research and clinical practice. Continued attention to these five topics, particularly preventive and pediatric-focused interventions, will be essential for further advancement in managing dental anxiety effectively.

## Conclusion

This comprehensive bibliometric study provided insights into the dynamic progression of dental anxiety research from 1991 to 2024. The analysis underscored an increased interest in the topic in recent years and significant thematic expansions and methodological complexity, particularly in the development of assessment tools and intervention strategies. A notable increase in multidisciplinary treatments, including cognitive-behavioral approaches, pharmacological aids, and innovative adjunctive therapies such as virtual reality and aromatherapy, characterized recent publications.

Furthermore, *pediatric dentistry* emerged prominently as a critical domain, emphasizing the necessity of early and tailored interventions to mitigate long-term dental anxiety. The evolving research underscores a shift towards multi-disciplinary, patient-centered practices, enhancing oral health outcomes and patient comfort. Future investigations should continue prioritizing interdisciplinary approaches, particularly those integrating psychological interventions and emerging technologies, to further improve patient management and preventive strategies. Continued focus on prevention and early evidence-based treatments for affected individuals in pediatric populations remains essential for addressing dental anxiety at its developmental stages, ultimately promoting lifelong positive attitudes toward dental care, thereby greatly improving oral health.

## Disclosures

### Conflicts of interest disclosure

The authors of this work declare that they have nothing to disclose.

### Availability of data and materials

Data generated and/or analyzed during the current study are available from the corresponding author on reasonable request.

### Funding sources

None.

### Artificial intelligence

No artificial intelligence has been used in any part of the analysis or writing.

## Data Availability

All relevant data are within the manuscript and its Supporting Information files.

## Acknowledgements

Not applicable

## Authors’ contributions

YH, RV, RS, and NC contributed to the conceptualization, and methodology. YH, RV contributed to data collection, data curation, and formal data analysis. YH, RS, and NC contributed and interpretation. RS and NC wrote the original draft, while YH and RV reviewed the manuscript. Finally, all authors read and revised the manuscript prior to submission.

## Notes

### Competing Interest Statement

The authors have declared no competing interest.

### Funding Statement

The author(s) received no specific funding for this work.

